# The impact of Short Tandem Repeats on grey matter brain imaging derived phenotypes in UK Biobank

**DOI:** 10.1101/2023.02.27.23286496

**Authors:** William Sproviero, Upamanyu Ghose, Laura M Winchester, Marco Fernandes, Danielle Newby, Daisy Sproviero, Najaf Amin, Bart Smets, Karen Y. He, Ekaterina A. Khramtsova, Parth Patel, Brice A. J. Sarver, Trevor Howe, Mary Helen Black, Cornelia van Duijn, Alejo Nevado-Holgado

## Abstract

We performed a genome-wide association study of 143,067 highly polymorphic short tandem repeats (STRs) with MRI brain grey matter volumes (GMVs) on 10,702 UK Biobank (UKB) participants, including 8,751 in the discovery stage and 1,701 in the replication analysis. STRs’ repeat lengths were estimated from the UKB whole-genome sequencing data using Expansion Hunter software.

A total of 262 STRs reached genome-wide significance in the analyses of the autosomal and sex chromosomes’ (*P* = 6.9 × 10^−8^) in association with MRI -GMVs. Replication in a second batch extraction in the UKB and linkage disequilibrium (LD) analyses confirmed 12 associations of five STRs with hippocampal, intra-calcarine cortex, and cerebellum volumes with no evidence of single nucleotide polymorphisms (SNP) in LD detected in the surrounding DNA regions. Our study highlights the importance of STR variants involved in the genetic architecture of grey matter volumes.

## Introduction

Short tandem repeats (STRs) are variable length size sequences of short repeated DNA units (2-20 bp) and represent 3-6% of the entire genome.^1,2^ The increasing use of sophisticated bioinformatics tools and DNA/RNA sequencing technologies are opening the possibility of *in-silico* detection of STRs^3–6^ from whole genome sequences (WGS) data to perform genome-wide association analyses of STR variation at a population scale. These STRs are often multiallelic and seldomly in linkage disequilibrium with neighbouring SNPs.^5^ A proportion of these STRs are highly polymorphic, vary in length across individuals, and are associated to diseases.

In terms of disease, nearly 30 hereditary disorders are caused by expansions of specific STRs (e.g., spinocerebellar ataxias, Huntington Disease)^7^. It is hypothesised that STRs may contribute to explaining a portion of the missing heritability of common complex disorders (i.e., those caused by a combination of environmental and genetic factors), which is a frequently observed discrepancy between SNP-heritability and broad-sense heritability estimated by studies of siblings and relatives.^8^ In autism cases, for example, it was recently found in two studies that 54 STRs were associated at genome-wide significance level,^2,9^ and further 2,588 rare expanded STRs occurred in genes involved in nervous system development, cardiovascular system and muscle tissues.^2,9^ Other studies have implicated repetitive elements and structural variants in bipolar disorder^10^, schizophrenia^10,11^ and resistance to malaria.^12^

Recently, a phenome-wide association study was carried out on Exome sequencing data in the UK Biobank (UKB) to characterize the effect of de novo tandem repeats in human traits.^13^

Grey matter volumes (GMVs) and their longitudinal changes are very relevant to several psychiatric and neurological disorders. They have been shown to predict early stages of disease progression in neurodegenerative disorders (e.g., Alzheimer’s disease^14^, multiple sclerosis (MS)^15^), detecting brain atrophy related to pathological processes of disease. Major GMV changes are well-described in psychiatric disorders such as major depression disorder^16^, bipolar disorder^16^ and schizophrenia^17^. In the present study, we performed a genome-wide association study of highly polymorphic STRs (STR-GWAS) and MRI brain GMV phenotypes to assert the role of common STRs as genetic contributors to brain volumetric phenotypes using the first WGS data release of the UKB.

## Results

### Population Descriptive

We included unrelated European individuals with brain imaging phenotypes (Supplementary Table 1) and WGS data for a total of 10,702 participants. In the discovery analysis we included 8,751 individuals (mean age, 64.2 years; 52.5% women) and 1,701 individuals in the replication analysis (approximate mean age, 64.2 years; 54.6% women), both selected randomly as two separate extraction batches. A flow chart of all the analysis stages is reported in Fig. 1. The discovery and the replication cohorts had similar mean age and sex ratios. Descriptive statistics are reported in Supplementary Table 2. As a sensitivity analysis, 2,176 participants with prevalent neurodegenerative and related conditions that may influence brain volumes were excluded from the discovery analysis (see Supplementary methods and Supplementary Table 3) resulting in a cohort of 6,575 individuals. Brain MRI GMV image-Derived phenotypes (IDPs) in discovery and sensitivity or discovery and replication did not differ (Supplementary Table 4). Sensitivity analyses differed from discovery phase, which could potentially be attributable to a reduced sample size (Supplementary Table 5).

**Fig. 1.**
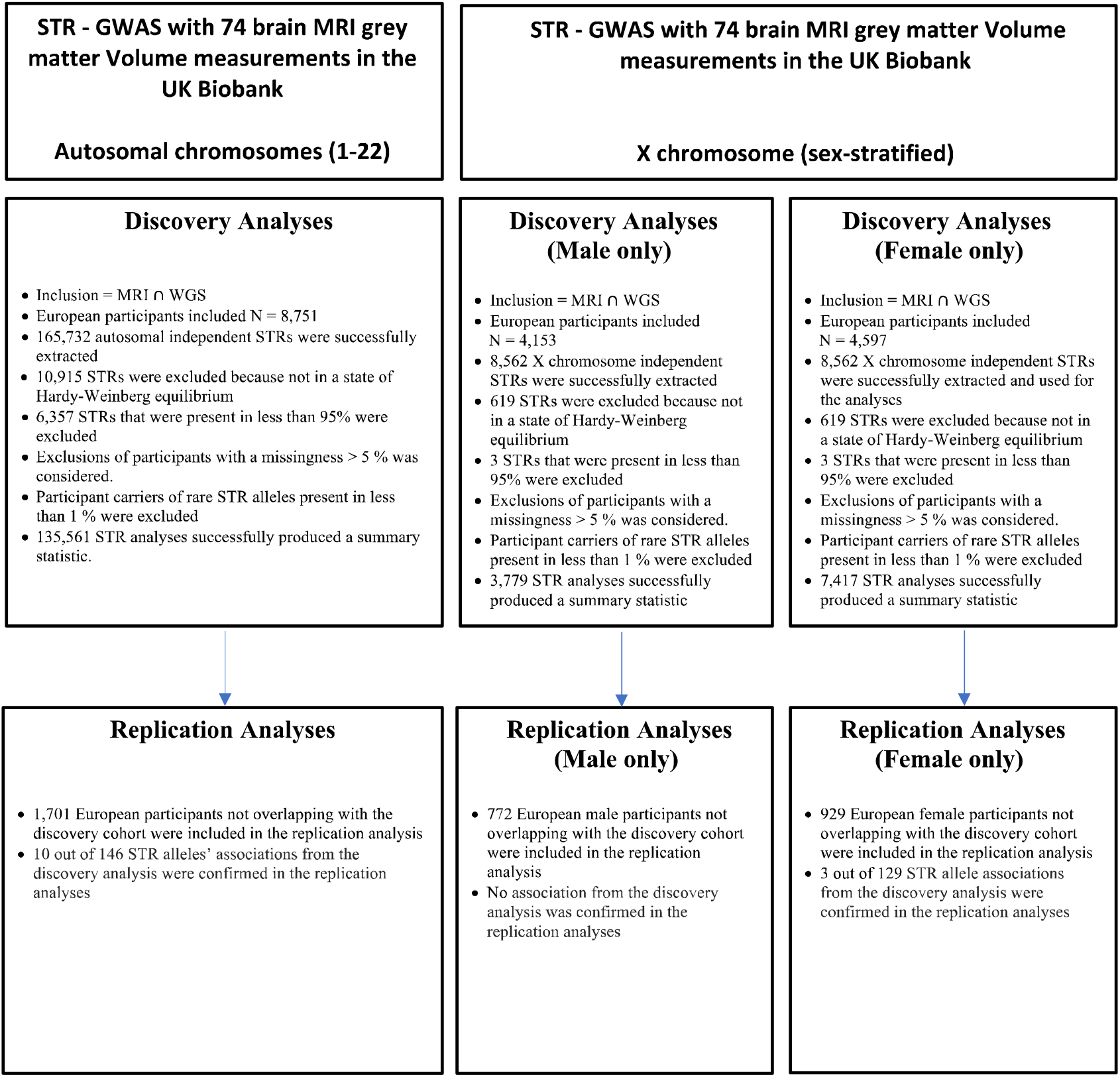
Study Analysis Flow Chart. A Weighted Least Squares Regression Model was performed in all analyses. A linear model’s outlier exclusion was applied at each stage using a Hat-value threshold > 0.5 followed by a heteroscedasticity test (*P* < 0.01). If a non-uniform distribution of the variance was detected, a robust estimation of the standard error was performed for the relevant STR tests. ⋂ = Intersection. Each STR analysis was conducted including a short and a long allele.

**Fig. 2.**
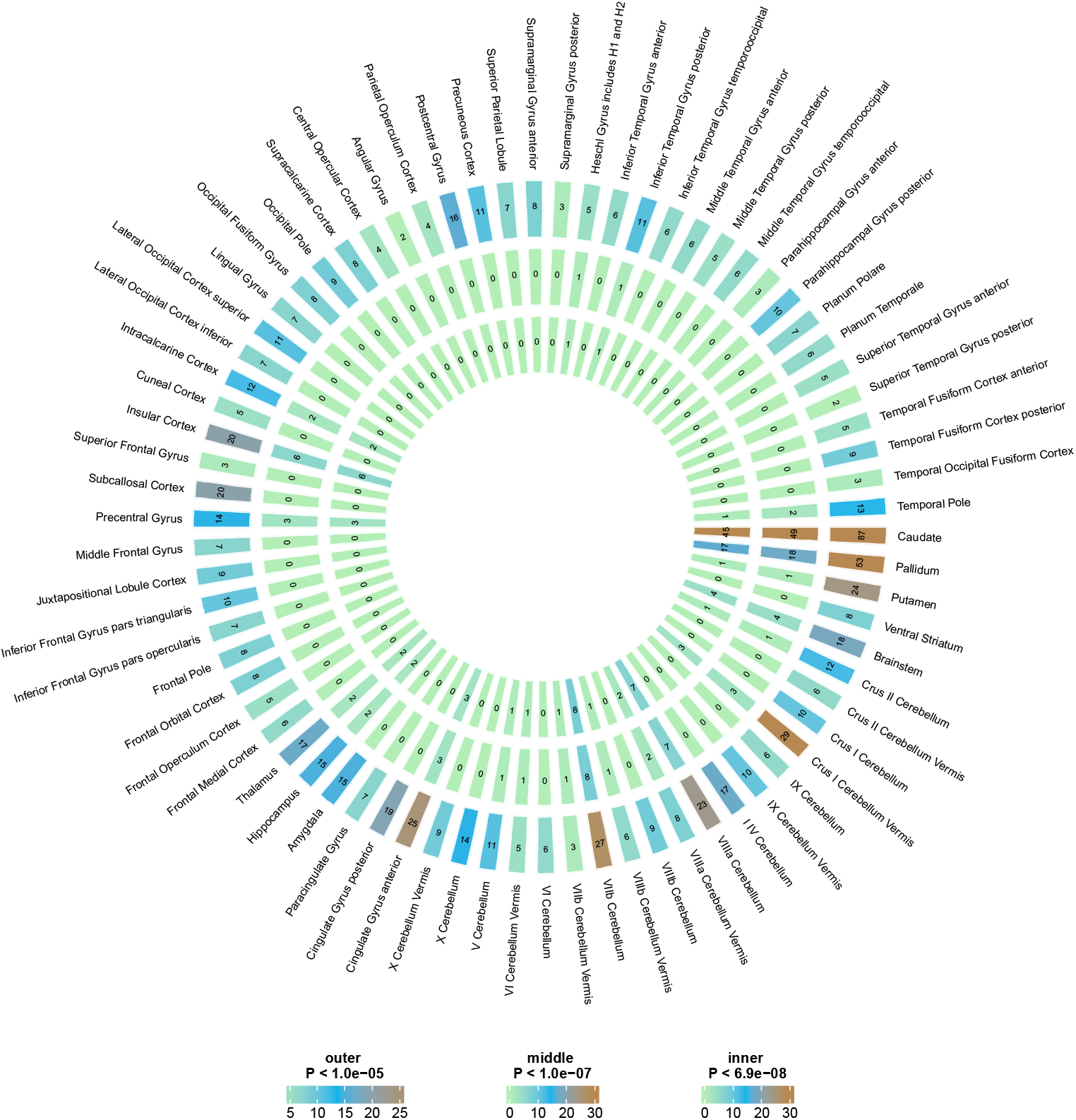
The count of significant STR associations with brain region volumes discovered at different significance levels. The outer layer represents the count of STRs for each brain region with P < 1×10^−5^, the middle layer represents the count of STRs with P < 1×10^−7^, and the inner layer represents the count of STRs with P < 6.9×10^−8^. The colour of the heatmap cells has been clipped at a maximum value of 29 to prevent the plot from being skewed by the regions with a large number of significant STRs. The plot was generated using the circlize^76^ package in R.

### Heterozygosity Comparison between UKB and 1000 Genome European populations

We found that the heterozygosity rates for the Expansion Hunter^3^ v5.0.0 (see URL) STR calls were highly similar between the UKB British cohort and the other five European 1000 Genome dataset sub-populations. The regression slope was 0.82 (adjusted R^2^ = 0.74, *P*-value < 0.0001) between UKB and GBR, and ∼0.80 (adjusted R^2^ ∼ 0.74, *P*-value < 0.0001) when compared to the other populations based on comparison of 147,310 STRs (Supplementary Table 6).

### Discovery Analyses: GWAS associations of 74 brain MRI GMVs

A total of 6,360 STRs were excluded from the analyses due to low STR calling rate -less than 95 percent - and 11,534 STRs were removed because distributions were not in Hardy-Weinberg equilibrium. After removing STRs with heterozygosity < 0.1, a total of 135,561 autosomal and 7,508 chrX’ STRs were successfully analysed in association with 74 brain GMVs.

There was no evidence for genomic inflation with λ_GC_ for each STR-GWAS ranging between 0.98 -1.03 (Supplementary Table 7). A total of 97 autosomal STRs, 50 chrX STR alleles for the male analysis, and 115 chrX STR regions for the female analysis were significantly associated with 30 brain MRI volumes. Six pairs of STRs, located in the same genomic regions, were associated with the caudate GMV in the male and female analyses. Six pairs of STRs, localised in the same genomic regions, were associated with the basal ganglia (caudate, putamen, pallidum GMVs) in the male and female analyses. All significant associations passed Bonferroni multiple testing correction of 6.9 × 10^−8^, i.e., 0.05/(143,067 STRs x 5 allele terms). Statistically significant autosomal and chrX analyses’ results are reported in supplementary Table 8-9, respectively.

### Replication of discovery lead STRs

For replication analysis we randomly selected, *a priori*, a second batch of extraction of 1,701 samples from the 10,702 UKB participants with both MRI volume and WGS data information. We used this subset to test the 97 autosomal and 50 + 115 chrX STRs (male and female associations) that passed Bonferroni correction in the discovery phase of the study in 30 brain volumes’ analyses. Results are reported in supplementary Table 8-9.

Six STR regions were significant with consistent direction of effect in the replication analysis (Table 1A-B). An STR (chr15:39340533-39340549, AC) located in the *AC013652*.*1* gene was associated with the precentral gyrus volume. An STR (chr14:57129041-57129077, TTA) located in the *OTX2-AS1* gene region was associated both to the Crus-II and VIIb cerebellum regions’ volumes. Another STR (chr2:156342362-156342390, CCCCCGG), located in a CpG island of the promoter region of the *NR4A2* gene, was associated with the hippocampus region volume. Two neighbouring STRs (chr14:59173671-59173695, AAAT; chr14:59140004-59140022, TG) ∼33.5 kB from each other (located in the upstream area close to the *DAAM1* gene) were both associated to the intra-calcarine cortex volume. One STR (chr1:75553883-75553913, AGG) located in the *SLC44A5* gene was replicated in three cerebellum vermis volume measurements (VIIb, VIIIa, VIIIb).

**Table 1a.**
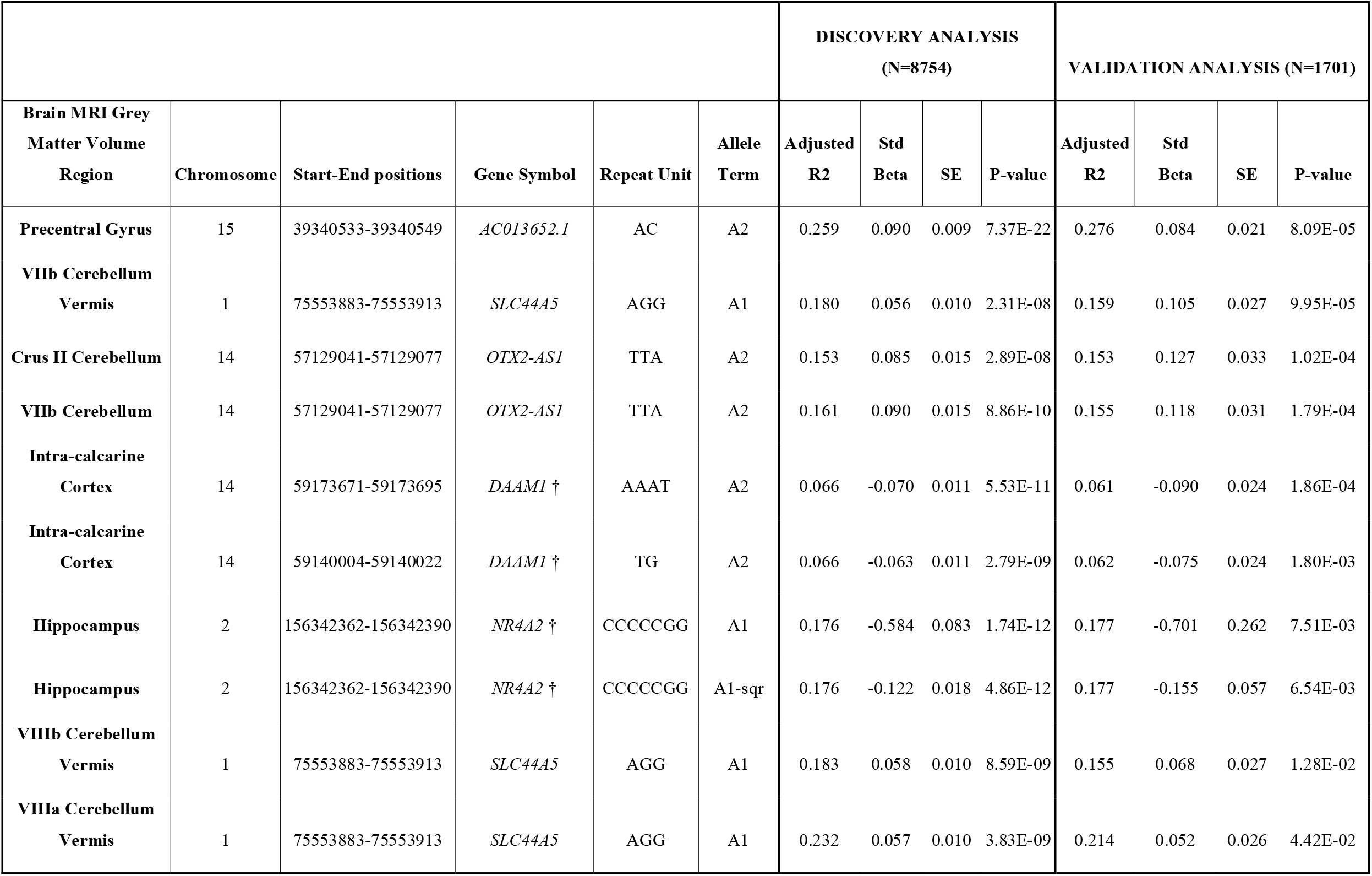
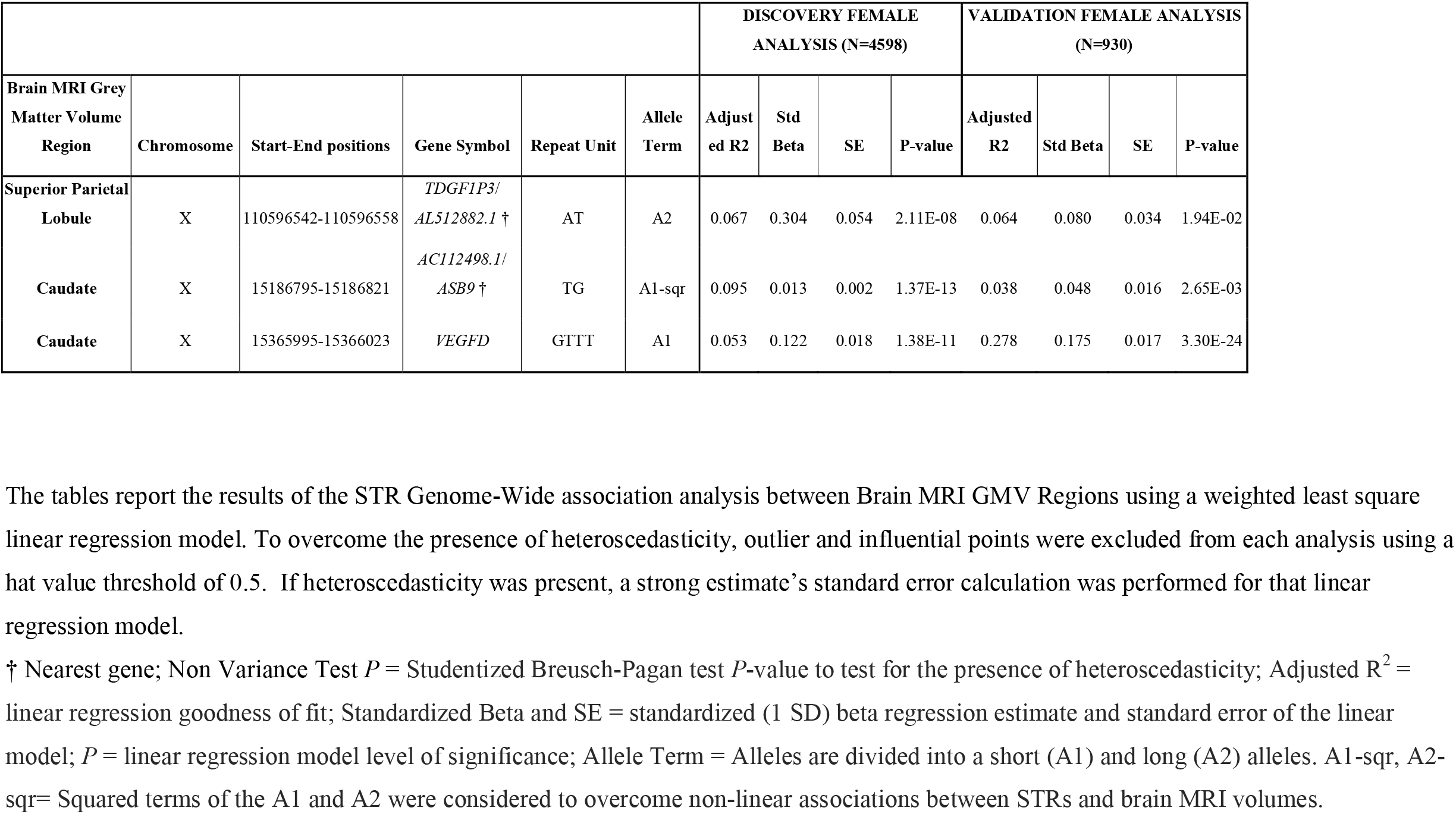

In the female-only analyses, we found three chrX STR regions that were confirmed in the replication analyses. Of the three significant chrX STRs, two were in association with the caudate [*AC112498*.*1/ASB9* (chrX:15186795-15186821, TG); *VEGFD* (chrX:15365995-15366023, GTTT)] and one with the superior parietal lobule [*TDGF1P3/AL512882*.*1* (chrX:110596542-110596558, AT)] volumes.

The replicated STRs were all included in DNA regions previously associated with GMVs and other traits in SNP-based GWAS. An overview of previous associations in the same gene regions is shown in Fig. 4.

### Linkage Disequilibrium analysis

A decay of LD is observed when comparing STR-SNP and SNP-SNP LD.^18,19^ Therefore, we performed an LD analysis between SNPs and STRs in the same region to highlight the importance of using STRs in addition to traditional SNP GWAS methodology. SNP-STR LD analysis was performed after phasing STR genotypes with phased SNP data from the European CEU and IBS population in the 1000 Genomes project.

Of the six autosomal and three chrX STR regions that passed statistical significance in the replication cohort, the following STRs did not report any tagging SNPs in LD (R^2^ < 0.8): one STR in the *OTX2-AS1* gene region (chr14:57129041-57129077, TTA); two STRs located in the upstream genomic region of the *DAAM1* gene (chr14:59173671-59173695, AAAT and chr14:59140004-59140022, TG); one STR in the *NR4A2* gene region (chr2:156342362-156342390, CCCCCGG); one STR in the *SLC44A5* gene (chr1:75553883-75553913, AGG); one STR in the *AC112498*.*1*/*ASB9* region (chrX:15186795-15186821, TG).

One autosomal association present in the *AC013652*.*1* gene region (chr15:39340533-39340549, AC); and two STRs present on the chrX were in LD with SNPs in each specific surrounding region: the *TDGF1P3*/*AL512882*.*1* region (chrX: 110596542-110596558, AT) and the *VEGFD* gene (chrX:15365995-15366023, GTTT).

A strong correlation was present between two STRs localised in the upstream genomic region of the *DAAM1* gene (LD R^2^ = 0.93, CEU population; LD R^2^ = 0.80, IBS population).

A table with a list of the STR – SNP and STR – STR LD R^2^ is reported in the Supplementary Table 10.

LD SNPs rs-numbers were queried in the GWAS catalog, showing no previous association with brain measurements but one SNP (rs7182018) located in the surrounding region of the *AC013652*.*1* gene. The rs7182018 was previously found in association with increased GMV in the central sulcus^21^ and with the aparc-Desikan rh pars triangularis volume^22^.

### Autosomal chromosomes: gene concordance between SNP-GWAS and STR-GWAS results

To understand the relevance of the significant STRs and how they associate with already known genetic markers, we searched for an association between the 97 autosomal significant STR’s genomic regions (gene or neighbour genes) and traits reported in the NHGRI-EBI GWAS catalog.^22^ We found that 66 of the 97 significant STR regions (66.6%) were located in genomic regions which were reported as associations with other traits in previous SNP-based GWAS (Supplementary Table 11).

Of these 66 STR, we found that seven STR genomic regions [(a) chr1: 50961014-50961046, TTTA; (b) chr5:113093469-113093491, TC; (c) chr20: 53752297-53752321, GT; (d) chr1:75553883-75553913, AGG; (e) chr2:32440751-32440778, TTA; (f) chr5:66344987-66345007, TG; (g) chr14:59173671-59173695, AAAT and chr14:59140004-59140022, TG] were previously reported in GWAS of brain GMVs, including (a) *CDKN2C* gene region associated with amygdala and cerebellum exterior volumes^23^; (b) *MCC* gene region associated to cerebellar vermal lobules VIII-X volumes^23^; (c) *RNU7-14P* gene associated to inferior parietal volume^23,24^; (d) *SLC44A5* gene region associated to cerebellar vermal lobules I-V, cerebellar vermal lobules VI-VII and cerebellar vermal lobules VIII-X^23^; (e) *BIRC6* associated with pallidum volume^25^; (f) *LINC02229* gene region associated to nucleus accumbens volume^25^; (g) *LINC01500/DAAM1* region associated to pericalcarine, precuneus, cuneus, supramarginal, lateral occipital, lingual, inferior parietal, fusiform, entorhinal cortex^23^ and occipital volumes^26^.

In addition, we compared the overlap of our STR regions results with published results obtained from GWAS of 3,144 IDPs from a subset of UKB publications^27–30^. A total of 14 STR regions localised in genomic regions associated with IDPs (Supplementary Table 11). Six of these 14 regions were in association with brain volumes (*BIRC6* (chr2:32440751-32440778, TTA), *CDKN2C* (chr1:50961014-50961046, TTTA), *CLINT1* (chr5:157837411-157837439, AC), *DAAM1* (chr14:59173671-59173695, AAAT and chr14:59140004-59140022, TG), *MCC* (chr5:113093469-113093491, TC), *SLC44A5* (chr1:75553883-75553913, AGG)].

Gene concordance between STR-GWAS and SNP-GWAS results associated to traits other than brain volumes are reported in the supplementary results and in supplementary Table 11.

### X chromosome: gene concordance between SNP-GWAS and STR-GWAS results

The same NHGRI-EBI GWAS catalog^22^ search performed on autosomal significant STRs was applied on significant chrX STR regions. Of the 50 unique STR regions associated with MRI volumes in male participants, 18 STRs were located in genomic regions which were reported as associations with other traits in previous SNP-based GWAS associations (Supplementary Results; Supplementary Table 12a).

Of the 115 unique STR regions in association with MRI volumes in female participants, 30 STRs were located in genomic regions which were reported as associations with other traits in previous SNP-based GWAS associations (Supplementary Results; Supplementary Table 12b).

There were six gene regions [*GPC3* (chrX:133923131-133923151, AC; chrX:133882535-133882559, TG)], *ARMCX4* [(chrX:101479304-101479354, CA; chrX:101482388-101482408, TTCTT)], *AL157778*.*1* [(chrX:98610075-98610097, GA; chrX:98610075-98610097, GA; chrX:98738577-98738601, CA)], *DIAPH2* [(chrX:97483399-97483413, AG; chrX:97417375-97417397, AC)], *EDA* [(chrX:69674102-69674146, CTAT; chrX:69719715-69719743, AC)], *USP11* [(chrX:47243264-47243284, AAAT; chrX:47263009-47263044, TTTTG)] common between the male and female results (Supplementary Table 9).

A total of 10 STRs from the male analyses and 11 STRs from the female analyses were localised in genomic regions identified in IDP GWAS associations (Supplementary Tables 11a-b).^27–30^ Eight [*ARX* (chrX:25028556-25028577, TTA), *ARMCX4* (chrX:101479304-101479354, CA), *AL590065*.*1*/*CFAP47* (chrX:35882607-35882631, ATC), *IL1RAPL2* (chrX:104876718-104876745, TTA), *MECP2* (chrX:154037143-154037185, AT), *PLAC1* (chrX:134560494-134560512, GT), *SSU72P1*/*HTR2C* (chrX:114530672-114530690, CA), *Z75741*.*1*/*ACTRT1* (chrX:127807475-127807499, AAAT)] out of 10 regions for the male, and three (*MID1* (chrX:10761242-10761262, AC); *ATP11C*/*CXorf66* (chrX:139954713-139954734, AAC); *ARMCX4* (chrX:101482388-101482408, TTCTT)] out of 11 in the female analyses were in association with brain volumes.

### Pathway Enrichment

We ran a gene set enrichment analysis (GSEA) across all 74 brain volume STR – GWAS genes (Supplementary Table 13-14). A total of 44 pathways were reported (Fig. 3A). The axon guidance pathway was enriched in greater than 60% of tested brain areas. Eight pathways were enriched between 40-60% brain areas. Those pathways are calcium signalling, focal adhesion, neuroactive ligand receptor interaction, pathways in cancer, ErbB signalling pathway, long-term depression and arrhythmogenic right ventricular cardiomyopathy (ARVC).

**Fig. 3.**
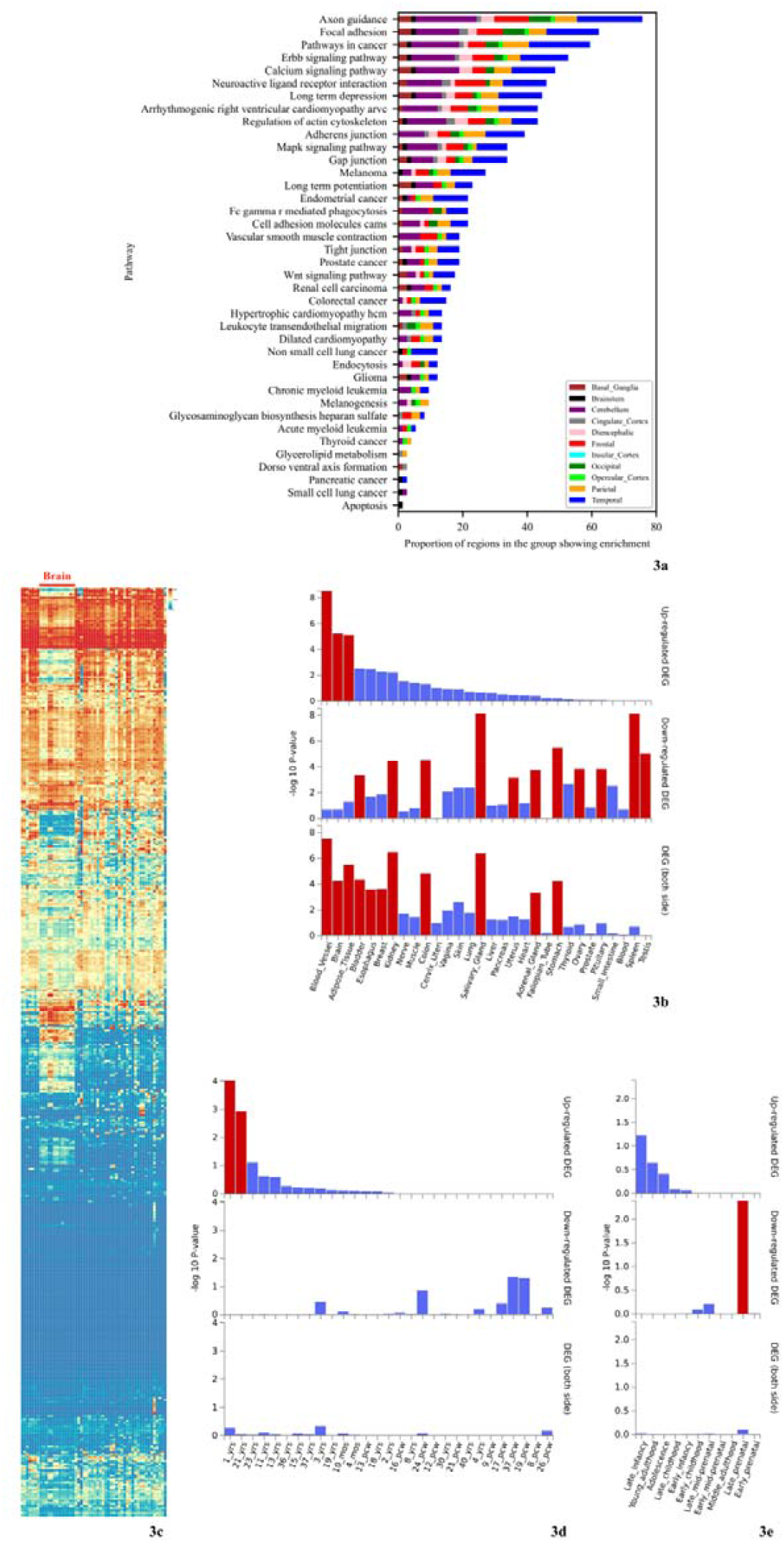
Pathway Enrichment and FUMA GENE2FUNC analyses: genes below nominal significance across all brain regions revealed an enrichment in brain involved pathways and a specific up-regulation in the brain in the early development stage. (a) A stacked bar plot showing a proportion of brain regions within a brain area group used to show the enrichment for a given pathway across all tested brain volumes. The proportion of brain regions in each broader brain group is represented by the different colours. The contribution of a brain group to the stacked bar plot is proportional to the number of brain regions present within that brain group. Any pathway that was not enriched in ≥ 30% of the brain regions of at least one brain group, was excluded from the plot. (b) Heat map demonstrating expression pattern of 528 human genes that mapped in any STR region that reached statistical significance of *P* < 1 × 10^−5^. A red bar below the 13 columns represents brain regions showing brain differential mRNA expression compared to other tissues. Heat map was generated using the GENE2FUNC platform. (c) Tissue specific analysis showing gene expression up-regulation in the blood vessel, brain, and adipose tissue and down-regulation in the salivary gland and spleen. Each tissue was compared to all other 29 tissue types, with significant differentially expressed gene (DEG) sets using a two-sided t-test (*Pbon*□<□0.05 and absolute fold change□≥□0.58) highlighted in red as described on the GENE2FUNC platform. (d) Brain gene expression comparison of any particular age among 29 different age grouti, ranging from 8 postconceptional weeks to 40 years old, with all other ages. Significant up-regulated expressed gene sets at one year age group are shown in red after Bonferroni correction with corrected *P* < 0.05 and absolute fold change ≥ 0.58 using the standard two-sided t-test described on the GENE2FUNC platform. (e) Brain gene expression comparison of any particular group among 11 different developmental stage groups, from early prenatal to middle adulthood, with all other stages. Significant up-regulated expressed gene sets at one developmental stage group is shown in red after Bonferroni correction with corrected *P* < 0.05 and absolute fold change ≥ 0.58 using the standard two-sided t-test described on the GENE2FUNC platform.

**Fig. 4:**
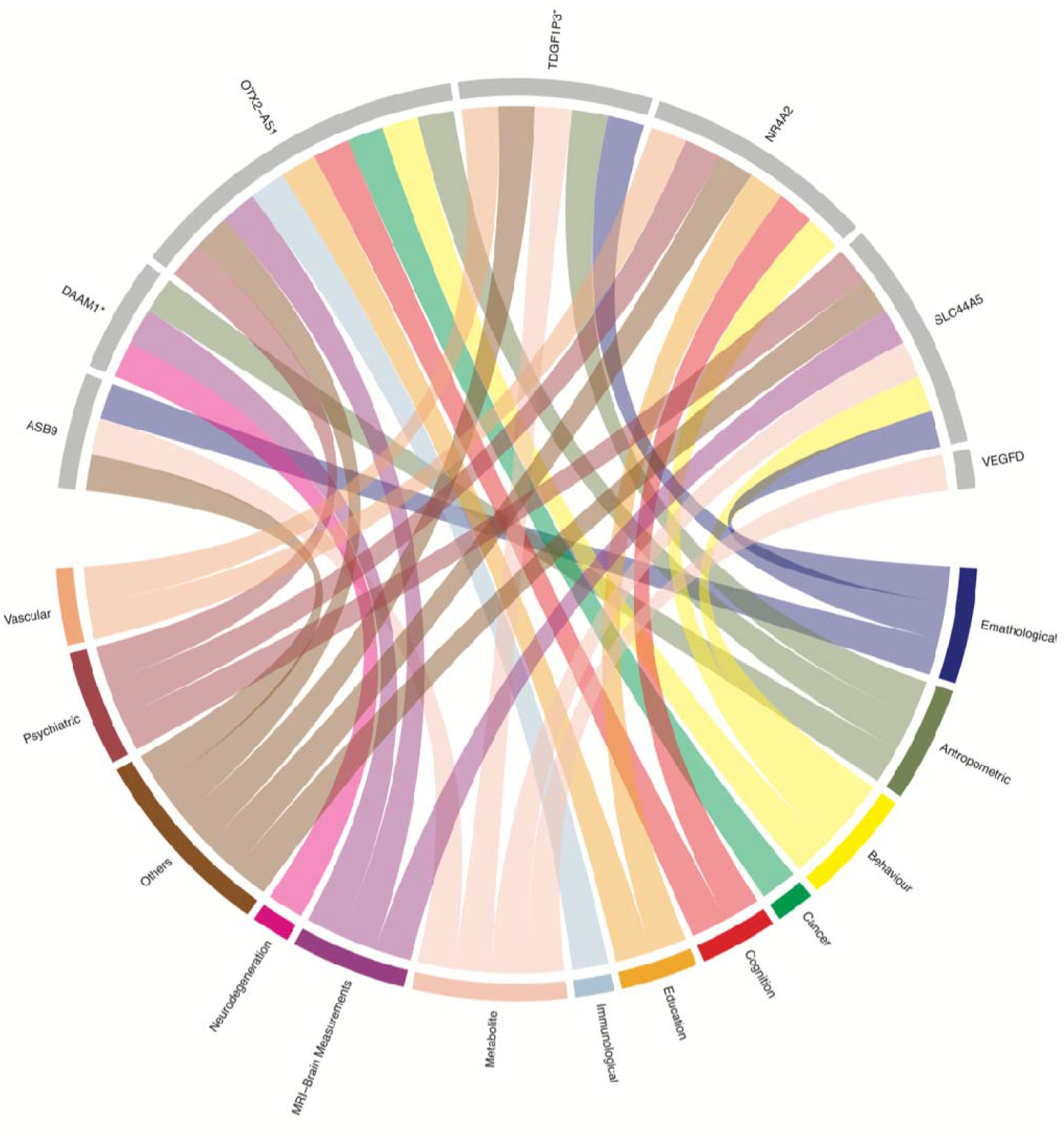
GWAS catalog disease/trait annotation of genes in which STRs were found associated with MRI-Brain GMVs’ measurements. Plot showing replicated STR mapped gene regions of interest that were previously associated with different traits/disorders in SNP-based GWAS. GWAS catalog^21^ web-tool was used to retrieve genes’ published association. Each major trait or disorder was assigned with different colours which highlight the link to the replicated STR mapped gene regions. The plot was generated using the circlize^74^ package in R. * = Nearest gene to the STR in association with brain MRI GMVs.

Only four pathways were enriched in around 40% of tested brain areas, and these were involved in cell organisation — adherens junction, regulation of actin cytoskeleton, gap junction and tight junction. Eight pathways were enriched in a proportion of 20-40%, among which included the relevant pathways MAPK signalling, long-term potentiation and Wnt signalling pathway.

### STR regions annotated genes (*P* < 1 × 10^−5^) across all brain analyses show a global up-regulation in the brain

We applied the publicly available GENE2FUNC function of the FUMA GWAS platform^30^ to a list of 528 genes (with a unique Ensembl ID) obtained mapping STR regions to genes that reached a statistical significance of P < 1 × 10^−5^ in any of the 74 GMV performed analyses (Supplementary Table 15). In the broader set of GTEx 30 tissue types, the 528 genes set showed significant up-regulation in the blood vessel (*P*_*adj*_ = 8.97 ×10^−8^), the brain (*P*_*adj*_ = 5.93 × 10^−6^) and the adipose tissue (*P*_*adj*_ = 7.60 × 10^−6^). Results are reported in Figure 3B and in the Supplementary Table 16. A visualization of the differential gene expression in 13 brain tissue regions amongst the GTEx 54 tissue types is reported in Fig. 3C and Supplementary Table 17).

Further over-representation analysis of the 528 gene list when comparing brain gene expression of 29 different age groups, revealed a higher mRNA expression in the human brain of the one-year age group (*P*_*adj*_ *=* 2.73 × 10^−3^) and of the 21-year age group (*P*_*adj*_ *=* 3.47 × 10^−2^) (Fig. 3D) (Supplementary Table 18).

A down-regulation of the gene set was observed in the prenatal developmental stage (*P*_*adj*_ *=* 4.5 × 10^−2^) compared with other 10 defined developmental stages from early prenatal to middle adulthood (Fig. 3E) (Supplementary Table 19).

## Discussion

We have evaluated the impact of 143,067 highly polymorphic STRs across the whole genome on 74 FAST MRI GMVs using data for 10,702 UKB participants of European origin subdivided in a discovery and a replication phase, and chrX analyses stratified by sex. A total of 30 GMV analyses detected 146 STR alleles in the autosomal chromosomes and for the chrX 50 and 129 STR alleles in male and female analyses, respectively, that passed Bonferroni threshold (6.9 × 10^−8^) in the discovery stage. A great majority of the significant STR results reported in this study were associated with caudate and pallidum GMVs (Fig. 2).

The results in the discovery phase provided evidence of an over-representation of STRs present either within gene boundaries or near genes whose mRNA expression is up-regulated in the brain, blood vessel and adipose tissue. This suggests the importance of vascularization of the brain for the development in synchrony and in a mutually dependent manner with the central nervous system^32^. Moreover, it supports evidence of cross-talks between the brain and distinct adipose tissue depots to maintain energy balance^33^.

We detected a down-regulation of STR gene regions in the late prenatal developmental stage and an up-regulation in brains, respectively, highlighting a possible key role of STRs in regulating genes that are important, after birth, during the growth of axons and dendrites with dendritic spines (neuritogenesis) during the first two years of life^34^ and, also, important in the maturation of the prefrontal cortex that continues evolving through the adolescence until the early 20s^35,36^. A pathway enrichment analysis using the results in the discovery phase, further, confirmed a major representation of genes across all brain regions that were important in neuron specific biological pathways, such as axon guidance, which is of main importance for neuron connections^37,38^.

All STRs confirmed in the replication stage were present in gene regions characterised by a great level of pleiotropy as shown in Fig. 4. These gene regions of interest were previously associated in SNP-based GWAS with different traits/disorders. Three of these genes (*SLC44A5, DAAM1* and *OTX2-AS1*) were found not only associated with MRI brain measurements (e.g., GMV).

The *SLC44A5* encodes a solute carrier protein, which is involved in metabolism of lipids and lipoproteins, and SNPs in this gene were found in association with cognitive impairment, neuroticism, depressive symptoms (Supplementary Table 11) and with early brain injury^39^. Our results confirm a finding that this gene may play a role in the genetic architecture of the cerebellum vermal lobules.

The Dishevelled-Associated Activator of Morphogenesis or *DAAM1* gene is a key component in the planar-cell-polarity signaling pathway^40^ and it is implicated in the elongation of new actin filaments and regulation of cell growth^41^. Previous findings reported an association of SNPs in the *DAAM1* region with neurofibrillary tangles and Alzheimer’s Disease (Supplementary Table 11). Polymorphisms in the *DAAM1* gene were also associated with a structure-predicted functional connectivity of the human visual cortex (Supplementary Table 11). Our finding confirms the association of two STRs present in the upstream region of this gene with the intra-calcarine cortex GMV.

The *OTX2* Antisense RNA 1 or *OTX2-AS1* is a long non-coding RNA (lncRNA) ^42^. SNPs in this region were previously found associated with educational attainment, mathematical ability, depression and neuroticism (Supplementary Table 11). Little is known about the role of this lncRNA. However, the attached neighbour gene *OTX2* is important in controlling the development of several neuronal populations in the midbrain and cerebellum during the neurogenesis^42^ and previous findings suggest that it may antagonize vulnerability to the Parkinsonian toxin MPTP^43^. Our findings confirm an association of one STR in the *OTX2*/*OTX2-AS1* region with the cerebellum GMV.

The *NR4A2* gene is a novel gene that has never been associated with GMVs before, but SNPs in this region were previously associated also with educational attainment, mathematical ability, and cognitive traits, as well as depression and neuroticism (Supplementary Table 11). The *NR4A* family of nuclear receptors, to which *NR4A2* belongs, is important in the hippocampal synaptic plasticity and the memory processes^44^. The *NR4A2* gene is a regulator of the midbrain dopaminergic neurons differentiation, maintenance and survival.^45^ Mutations and deletions in this gene were detected in both sporadic^46^ and familial^47^ patients affected by Parkinson’s Disease (PD) and of patients affected by autism^48^ and schizophrenia^49^. Moreover, a dysregulation of *NR4A2* gene expression was reported in post-mortem brain of patients affected by Alzheimer’s Disease (AD)^50^. Our results highlight a deeper meaning of one STR located in a CpG island of the promoter region of *NR4A2* in regulating the genetic architecture of the hippocampal region.

The replication of gene regions previously associated with brain GMVs discovered in SNP-based GWAS and associated with STRs in the same genomic region could suggest the presence of LD between STR reported in this study and surrounding SNPs. Five STRs localised in the *SLC44A5, DAAM1, OTX2-AS1* and *NR4A2* regions, that had concordant direction of effect through all stages of the study, were not in LD with any SNP in a surrounding DNA area of ± 50 kb. This highlights the possibility of a SNP independent role played by these STRs influencing the brain regions for which an association was found in this study. Even if the STR-SNP LD was weak in these regions, the two STRs localised in the upstream area outside the *DAAM1* gene boundaries were in LD and may imply a deeper biological meaning in association with the calcarine cortex.

Besides the replicated STRs, it is not surprising that several significant regions found in our discovery analyses are novel loci mapping genes not previously associated with any GMVs in published SNP-based GWAS. This can be explained by an LD decay observed previously when comparing STR-SNP and SNP-SNP LD^18,19^, estimating the SNP-STR LD to be approximately half the SNP-SNP LD^5^.

## Limitations and strengths of the study

Highly polymorphic STRs have a pleiotropic effect that may have different biological implications according to the repeat length (e.g. Spinocerebellar ataxia^51^; ALS^52^). In our analyses, we addressed whether common STR lengths influence brain volumes in the larger population. Individuals harbouring STR alleles with MAF < 0.01 were excluded from all analyses. This practice eliminated known pathological and possible *de novo* STR expansions that have higher penetrance and that lower the burden threshold for specific traits or diseases. However, these STR expansions are rare and possibly associated with specific pathological phenotypes.

This study also has limitations. Our sample consists of British participants of European ancestry, meaning that the generalizability of our results depends on the replication of the association of STRs of interest with specific GMVs in Non-European ancestries. The European bias in the majority of SNPs-GWAS remains relatively unknown^53^, and the European bias of STR-based GWAS compared to other populations has still to be evaluated and will depend on the heterozygosity of the STRs of each ancestry group^5^.

Moreover, in order to minimize the possibility of false positive results, our stringent linear regression model pipeline may have filtered out important STR allele lengths that resulted in some STR *P*-values not surviving the multiple testing correction. This is evident in the pathway enrichment and in the tissue enrichment analyses, in which 74 GMV discovery analyses highlighted a major representation of STRs (*P* < 1×10^−5^) that mapped within gene regions which are important in neuron specific biological pathways and whose mRNA expression is up-regulated in the brain.

The exclusion of participants with neurodegenerative or psychiatric conditions was performed as a sensitivity analysis for the significant STRs after the discovery stage to determine the influence of such individuals on the general performance of our model and influence on the results. A power calculation evidenced a drop in statistical power in the sensitivity analysis but 92 out of 146 STR alleles from the discovery analysis showed an association at a nominally significant level, suggesting a minor or no difference of results between the two analyses. We still might have missed conditions with long prodromal period (e.g. AD; Vascular Dementia)^54^, however, we do expect the signal produced by these participants to be taken care of by our strict statistical model pipeline which identified and excluded exceptional influential data points (see Methods section).

STR somatic instability is an important consideration in the measurement of repeat length. This instability is expansion-biased, age-dependent and tissue-specific. This implies that the number of repeats measured will be dependent: on the age at which the DNA sample was obtained; on a form of heterogeneity in the STR length, defined as mosaicism, that may be present across tissues of the same individual if the repeat has increased in length over generations. However, in the case of known expanded STRs (e.g. myotonic dystrophy type 1 (DM1)), it was shown that increases in the allele length or degree of somatic instability is negligible if the STR length did not reach the expanded length size.^55^

Moreover, in order to avoid any residual source of bias related to hidden relationships between age and STR lengths, age of the individuals at MRI visit was used as covariate in every statistical model used in this study. This was not the age when blood was drawn, however, the difference between the age at first MRI visit and the recruitment age was not greater than 9.14 ± 1.74 years across the included participants.

Tandem repeat tools available today make use of WGS data, hence, the accuracy of detection of STR lengths on both allele depends on the WGS depth, coverage and algorithm used for STR length size estimation.^56^ However, the STR detection software used in this study is a well-established method for estimating tandem repeats with a published record for detecting short and, as well, long STR allele length sizes^3^. Moreover, both UKB and 1000 Genomes sequence data used a WGS high coverage protocol which allowed a better accuracy of prediction. This is confirmed by the concordance in estimated heterozygosity of the STRs screened in this study by comparing the UKB and the 1000 Genomes European populations with an average regression slope of ∼ 0.82.

## Conclusion

In this study we explored for the first time the influence of STRs on the genetic architecture that underlies differences of brain volume phenotypes in the broad population using brain MRI measurements. We showed that STR-GWAS could complement standard SNP-GWAS association findings, highlighting the possibility of detecting association effects that may be only partially affected by LD association with other SNPs in the surrounding region.

Moreover, taking in consideration all brain GMV discovery analyses, genes that mapped STR DNA regions that reached a *P*-value < 1×10^−5^ significance showed an mRNA expression up-regulation in the brain, especially during the first year of life and 21 years of age.

Finally, this study opens the chance of detecting true causation by analysing highly polymorphic structures of the DNA which are characterized by a pleiotropic effect due to their variable repeat length intrinsic nature and provides a new insight to search for missing heritability^8^ in association with brain measurements and specific traits or diseases.

## Methods

### Participants Inclusion criteria

UKB participants were included if WGS data and FAST – MRI brain GMV information was available (N = 10,702) and only European participants (UKB field 21000) were considered in this study. A genetic relationship matrix (GRM) was built for these individuals using GCTA and a relationship threshold of 0.025 was applied to exclude related individuals. One person from each pair of related individuals was retained randomly.

Participants included in the study were randomly subdivided into a discovery (N = 8,751) and a replication (N = 1,701) stage as two separate extraction batches.

### Whole Genome Sequencing

The first release of UKB whole genome sequencing (WGS) data included 150,119 participants. DNA was sequenced to an average coverage of 32.5x and a minimum coverage of 23.5x per individual, using Illumina NovaSeq sequencing machines at deCODE Genetics (90,667 individuals) and the Wellcome Trust Sanger Institute (59,452 individuals). Individuals were pseudo-randomly selected from the set of UKB participants and divided between the two sequencing centres. After exclusion of duplicated samples and of samples of participants who drop out from the study, the final cohort of WGS data included 149,960 individuals. Details on WGS quality control procedure were described in a previous publication.^57^ Whole genome CRAM files (UKB field 23193) were used for the extraction of STRs.

### Genome-wide polymorphic STR catalog

A genome-wide STR catalog was used to perform association analyses between STR lengths and MRI brain GMVs. The algorithm applied to generate the STR catalog used in this study is described elsewhere^58^ and a summary of the method is reported in the Supplementary methods. The catalog comprises 174,293 highly polymorphic STRs which show varying lengths in all individuals. Their positions are relative to the genomic database hg38.^58^ The STR catalog includes ∼80% di- and tetra-nucleotide STRs. A total of 17% is represented by tri-nucleotide followed by penta- and hexa-nucleotide STRs. The remaining 3% is represented by STRs with a unit range between 7-24 base pairs (Supplementary Fig. 1). This is in agreement with previous data distribution of STRs across the genome.^5^

### STR detection and Quality Control

STRs were detected using ExpansionHunter (EH) software V5.0, an *in-silico* PCR-free method for estimation of STRs in WGS data aligned files (BAM/CRAM).^3^ EH has high accuracy for predicting both normal repeat lengths of less than 150 base pairs (WGS read length), and high abnormal expansions > 200 bp (e.g. C9ORF72 hexanucleotide; FXN trinucleotide; FMR1 trinucleotide)^3,59,60^ with an agreement between the gold standard fragment capillary analysis and in-silico prediction of 97.7 -99.0%.^3,59,60^ Samples were genotyped separately with default parameters: (i) excluding STR loci with coverage below 10 for diploid chromosomes and 5 for haploid chromosomes;(ii) on/off-target regions’ search for informative reads set to 1000 bp. EH outputs in the variant call format (VCF v4.1) were further merged across all samples.

The following downstream filters were also applied: (i) STRs which did not reach 95% call rate were excluded from all analyses; (ii) STRs whose frequencies did not meet the percentage of homozygous versus heterozygous calls expected under Hardy–Weinberg equilibrium (binomial two-sided *P* < 1 × 10^−9^) were excluded using the R package ‘HardyWeinberg’ 1.7.5^61^; (iii) a 5% sample missingness was also set as criterium of exclusion of participants.

In each STR model, a minor allele frequency (MAF) of 1% was applied to filter out rare alleles. STRs with heterozygosity < 0.1 were excluded a priori from all analyses.

*In-silico* STR genotype and filtering analyses were performed on DNAnexus RAP using a machine with 16 cores and 32 GB RAM.

### STR Heterozygosity Calculation

We calculated the heterozygosity (H_E_) of each STR as the aggregated frequency of each STR allele according to the formula 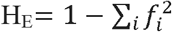, where f_i_ denotes the frequency of the i^th^ allele at the locus.^5^ Slopes and R^2^ values for STR heterozygosity comparisons between 1000 Genome European populations^62^ (CEU, FIN, GBR, IBS and TSI) and UKB were calculated using the linear regression function (lm) in R.

### (FAST) MRI -Brain GMVs

A list of all UKB data fields reporting FAST -MRI brain GMVs is included in the Supplementary Table 1. For 65 out of 74 GMV phenotypes, a left and a right brain hemisphere volume measure were reported in the UKB dataset and an average volume was calculated.

### GWAS -Linear Regression Model

Linear regression models were employed to assess the association of brain MRI GMVs and STR lengths at genome-wide level. Normalised MRI volumes were set as dependent variables (Supplementary Methods). STR lengths of both alleles (A1, A2), including the product term (A1 x A2) and the quadratic terms (A1^2^, A2^2^), were used as the main independent variables. Regression estimates were retained for each independent term per STR analysis.

Analyses for autosomal chromosomes were performed using all individuals, but separate analyses were performed for males and females for the chrX.

Linear regression models were adjusted for age (UKB field 21003, Instance 2), sex (UKB field 31), age^2^, age × sex, age^2^× sex; MRI centre (UKB field 54, Instance 2); scanner lateral brain position and scanner table position for T1-structural brain MRI [(X), UKB field 25756; (Y) UKB field 25757; (Z) UKB field 25758; scanner table position, UKB field 25759]; volumetric scaling from T1 head image to standard space factor needed to normalize for head size (UKB field 25000); and six genetic principal components correcting for population structure (UKB genetic PCAs). All continuous variables were standardized, and description is reported in supplementary methods.

From each model, we obtained the linear regression estimates, standard error, *P*-value and adjusted R^2^. In order to account for the effects of heteroscedasticity and influential points, we applied outlier exclusion criteria, followed by a weighted least squares (WLS) regression model to each STR analysis. In a least-squares fitting it is important to determine the influence of each value of each dependent variable on the estimated (i.e. fitted) values of the dependent variable. Therefore, a projection matrix known as the hat-matrix was used to identify exceptional influential data points.^63^ Hat-values range between zero and one, and a value > 0.5 was used as observation data point exclusion. After outliers were excluded, a WLS regression model, which incorporates the covariance matrix of errors in the model, was performed on each STR relative independent term.

The WLS regression is a robust method that controls for heteroscedastic distribution of the variance. A studentized Breusch-Pagan test (BPtest) was further performed to test any residual presence of heteroscedastic distribution of the variance in the linear model. BPtest *P* > 0.01 was considered as no presence of heteroscedasticity.

Heteroscedasticity does not affect linear regression estimates but bias only the linear regression standard error (SE). A last step of robust estimation of the SE was performed on the solely analyses with a BPtest *P* < 0.01.

All analyses were performed in the R environment (version 3.6) inside DNAnexus Research Analysis Platform (RAP) environment. The function “bptest”, as part of the R package “lmtest”, was used to perform the non-variance BPtest.

A Bonferroni threshold of 6.9 × 10^−8^ was requested for each test to reach genome-wide significance number of allele terms analysed — A1, A2, A1×A2, A1^2^, A2^2^ — (N=5) × number of STRs tested (N=143,067).

An inflation factor *λ*GC was calculated for each independent STR allele term’s *P*-value distribution.

In the final step, the statistically significant STR alleles that passed the Bonferroni threshold in the discovery phase were also analysed in a separate replication cohort. Replication analyses that reached a *P* < 0.05 were considered statistically significant.

Replication analyses were performed following the same procedure previously applied in the discovery phase. The “pwr.2p.test” function from the “pwr” R package was used.

### Gene concordance between SNP-GWAS and STR-GWAS results

For the detected STRs present either within gene boundaries or near genes we performed lookups in the NHGRI-EBI GWAS catalog^22^ (version 2022-07-30) to explore the previously reported SNP-based GWAS associations with the same or other traits. A significant *P* < 5 × 10^−8^ was considered to retain associated SNPs.

### Pathway enrichment analysis

We performed pathway enrichment with the autosomal STR-GWAS summary statistics using gene set enrichment analysis (GSEA).^64^ The Kyoto Encyclopedia of Genes (KEGG) annotations were used as the gene sets in the analysis. To implement GSEA, we used the R package “fgsea”.^65^ Using annotations from the SNPnexus web-based tool^66^, we mapped the autosomal STRs to the genes they lay within, or the nearest gene in case the STR was in a non-coding region. To obtain a gene-level statistic for fgsea, we selected the GWAS Z-score statistics of the STR with the maximum absolute GWAS statistic among all STRs mapped to a gene. The ranking statistic (“stats” parameter in the R function) for fgsea was set as the gene-level statistic obtained in the previous step, and the number of permutations (“nperm” parameter in the R function) was set to 1000. Furthermore, to better understand the enrichment pattern across the 74 brain regions, we retained only those pathways that were enriched (*P*_*adj*_ < 0.05) in at least one of the regions and generated a cluster-map using the *-log*_*10*_*(P*_*adj*_), where *P*_*adj*_ is the adjusted enrichment *P*-value of a region for a given pathway. The brain regions were clustered into 11 broader groups^67,68^ (Supplementary Table 12). Finally, to identify the most enriched pathways across the 11 groups, we generated a stacked bar plot of the proportion of brain regions within a group that showed enrichment for a given pathway. Any pathway that was not enriched in ≥ 30% of the brain regions of at least one group, was excluded from the bar plot to allow “clearer viewing”.

### Gene enrichment analyses

Autosomal STR regions whose *P*-values reached a statistical significance of less than 1×10^−5^ were annotated using SNPnexus web-based tool^66^. Gene IDs which overlapped or that were neighbours (the nearest gene was considered) of STR regions of interest were retained and used as input into GENE2FUNC function of FUMA^31^, an online tool which identified significantly up-or downregulated differentially expressed gene sets across human tissue types with a Bonferroni corrected *P* < 0.05.

GENE2FUNC utilises the GTEx v8^69^ data of 30 general tissue types, with all brain regions summarised as one tissue type, and the 54 specific tissue types that include 13 individual brain regions. Using the GENE2FUNC function, we further examined if these genes show relatively higher expression in BrainSpan^70^ when comparing (i) brain gene expression data from 11 defined developmental stages from early prenatal to middle adulthood, and (ii) brain gene expression of any particular age among 29 different age groups, ranging from 8 postconceptional weeks to 40 years old, with all other ages. Data is presented alphabetically, with differentially expressed gene sets shown in red after Bonferroni correction with corrected *P* < 0.05 and absolute fold change ≥ 0.58 using the standard two-sided t-test described on the GENE2FUNC function.

### Linkage Disequilibrium between STR - SNP

STRs that reached statistical significance in the replication phase were further investigated for the presence of SNPs in LD in the STR surrounding region of ± 50 kb. CRAM files from the 1000 Genome project of 57 trios (171 individuals) from CEU (Northern Europeans from Utah) and 50 trios (150 individuals) from IBS (Iberian populations in Spain) trios were downloaded from the 1000 Genome website repository^62^. In order to perform allele phasing between STRs and close surrounding SNPs, HipSTR^71^ v0.5 software was used to profile STRs in each sample by a joint multi-sample calling. HipSTR default parameters were used to call STR genotypes. Trio samples from the 1000 Genomes phase 3 SHAPEIT’s duoHMM phased SNPs^62^ matching samples used in the STR calling were provided as input to allow HipSTR to perform STR physical phasing when possible. HipSTR VCF outputs were filtered using the filter_vcf.py (for autosomal chromosomes) and filter_haploid_vcf.py (for sex chromosomes) scripts in the HipSTR package using suggested parameters (--min-call-qual 0.9 --max-call-flank-indel 0.15 --max- call-stutter 0.15). Beagle^72^ version 4.0 was used to phase each STR (up to ± 50 kb) separately using unphased STR, phased SNP genotypes and pedigree information. Beagle has been used in previous studies to phase multi-allelic STRs^73^. A pedigree file was provided to help LD estimation using family information. The methodology used is described in detail by Saini et al.^74^ Only unrelated parent samples were used from the phased STRs and SNPs outputs to estimate LD R^2^ between each STR and SNPs. Plink 1.9^75^ was used to obtain R^2^ estimates and R^2^ values > 0.8 were flagged as associated in LD.

IDs of SNPs in LD with STR regions were queried in the GWAS catalog to determine a previous association with other traits in other GWAS studies.

## Supporting information

Supplementary Methods and Results

## Data Availability

All data produced in the present study are available upon reasonable request to the authors.

## Data Availability

DNAnexus Research Analysis Platform (RAP), https://www.dnanexus.com/; 1000 Genome project phased variants (release version, 28/10/2022), https://ftp.1000genomes.ebi.ac.uk/vol1/ftp/data_collections/1000G_2504_high_coverage/working/20201028_3202_phased/; 1000 Genome project 30x GRCh38 whole genome sequencing CRAM files, FTP server; GCTA software, https://yanglab.westlake.edu.cn/software/gcta/#Overview; Plink 1.9, https://www.cog-genomics.org/plink/; SNPnexus annotation web-based tool v4, www.snp-nexus.org; Expansion Hunter v5.0 software https://github.com/Illumina/ExpansionHunter/releases/tag/v5.0.0; STR catalog https://github.com/Illumina/RepeatCatalogs/blob/master/hg38/variant_catalog.json; FUMA http://fuma.ctglab.nl/; HipSTR v0.5 software https://github.com/HipSTR-Tool/HipSTR; Brainspan, http://www.brainspan.org; GTEx v8 tissue mRNA expression, https://gtexportal.org.

Complete summary statistics of STRs in association with 74 GMV-IDPs are not publicly available but are available from the corresponding author on reasonable request.

## Code availability

Code used in this manuscript are not publicly available but are available from the corresponding author on reasonable request.

### Acknowledgment

We thank the UK Biobank participants and the UK Biobank team for their work in collecting, processing, and disseminating these data for analysis. This research was conducted using the UK Biobank Resource under the approved projects 52293 and 15181. Initial brain volume STR-GWAS was conducted using the application 52293. Bridging file between the two application was obtained with thanks to the UK Biobank and the Population Analytics research group, Research and Development, Janssen Pharmaceutical Companies of Johnson and Johnson. We thank Prof. Simon Lovestone for the helpful discussion on the project.

## Author Contribution

W.S. conceived and designed the study. W.S. had full access to the UK Biobank data in the study and take responsibility for the integrity of the data and the accuracy of the data analysis. W.S., L.W., B.S., B.A.J.S., E.A.K., P.P., K.Y.H., M.H.B., T.H., C.vD. and A.J.N-H. were involved in administrative, technical, or material support. W.S., U.G., M.F., D.N. and D.S. were involved in statistical analysis. All authors were involved in acquisition, analysis, or interpretation of data. WS, LW and UG were involved in drafting of the manuscript. W.S., U.G., L.W., M.F., D.N., D.S., B.S., B.A.J.S., E.K., K.H., P.P., M.H.B., T.H., C.vD. and A.J.N-H were involved in critical revision of the manuscript for important intellectual content. C.vD. and A.J.N-H. obtained funding. B.S., B.A.J.S., E.K., M.H.B., T.H., L.W., C.vD. and A.J.N-H supervised.

## Acronyms

GWAS; LD; MAF; SNP; STR; MRI; UKB

