## Supplementary Methods and Results for "The impact of Short Tandem Repeats on grey matter brain imaging derived phenotypes in UK Biobank"

**Genome-wide polymorphic STR catalog**

The software algorithm used to create the genome-wide short tandem repeat (STR) catalog is described elsewhere.^1^ In summary Dolzhenko et al^1^ inferred STRs from read alignments at population levels by detecting insertions and deletions from short WGS read alignment and extracting repeat-like sequences. Only repeats present in multiple samples were further processed to infer STR annotations.

A total of 174,293 STRs are present in the catalogue and include only highly polymorphic STRs and common STRs by requiring > 5% samples from the original cohort with alternative alleles^1^.

**Brain MRI GMV normalization**

Kolmogorov-Smirnov test was used to determine if GMVs were normally distributed. P-values of less than 0.05 were considered a sign of non-normal distribution. GMVs were normalized either using log-, square-root- or inverse-transformation. Kolmogorov-Smirnov test was calculated again to confirm the presence of a normal distribution (Data not reported) after transformation and the best normalization method was applied.

The following volumes were normalized using log-transformation: Parahippocampal Gyrus posterior division, Parietal Operculum Cortex, Planum Polare, Planum Temporale, Precentral Gyrus, Superior Parietal Lobule, Superior Temporal Gyrus anterior division, Superior Temporal Gyrus posterior division, Supramarginal Gyrus anterior division, Supramarginal Gyrus posterior division, Temporal Fusiform Cortex posterior division, Temporal Pole, V Cerebellum, X Cerebellum, X Cerebellum Vermis, Amygdala, Angular Gyrus, Central Opercular Cortex, Cingulate Gyrus anterior division, Cingulate Gyrus posterior division, Crus I Cerebellum, Cuneal Cortex, Frontal Operculum Cortex, Heschl Gyrus includes H1 and H2, IX Cerebellum, IX Cerebellum Vermis, I IV Cerebellum, Inferior Frontal Gyrus pars opercularis, Inferior Frontal Gyrus pars triangularis, Inferior Temporal Gyrus anterior division, Inferior Temporal Gyrus posterior division, Inferior Temporal Gyrus temporooccipital part, Intracalcarine Cortex, Middle Frontal Gyrus, Middle Temporal Gyrus anterior division, Middle Temporal Gyrus posterior division, Middle Temporal Gyrus temporooccipital part, Occipital Fusiform Gyrus, Occipital Pole, Pallidum, Paracingulate Gyrus and Parahippocampal Gyrus anterior division.

The following volumes were normalized using inverse-transformation: Postcentral Gyrus, Precuneous Cortex, Subcallosal Cortex, Superior Frontal Gyrus, Thalamus, Brainstem, Caudate, Frontal Medial Cortex, Frontal Orbital Cortex, Frontal Pole, Hippocampus, Insular Cortex, Lateral Occipital Cortex inferior division, Crus I Cerebellum Vermis, Lateral Occipital Cortex superior division and Lingual Gyrus.

The following volumes were normalized using square-root-transformation: Putamen, Supracalcarine Cortex, Temporal Fusiform Cortex anterior division, Temporal Occipital Fusiform Cortex, VIIIa Cerebellum, VIIIa Cerebellum Vermis, VIIIb Cerebellum, VIIIb Cerebellum Vermis, VIIb Cerebellum, VI Cerebellum, Ventral Striatum, Crus II Cerebellum, Crus II Cerebellum Vermis, Juxtapositional Lobule Cortex formerly and Supplementary Motor Cortex.

**Sensitivity Analysis - Diagnosis exclusions**

To address the possibility that associations between STRs and brain volumes might be driven by participants with prevalent neurological disorders, we excluded participants with these conditions.^2^ The UKB category of first occurrence of diagnosis (UKB category 1712) was used to exclude participants. Data variables reported under this category were generated including Read code information in the Primary Care data (category 3000); ICD-9 and ICD-10 codes in the Hospital Inpatient data (c[ategory 2000](https://biobank.ctsu.ox.ac.uk/crystal/label.cgi?id=2000)), ICD-10 codes in Death Register records ([Field 40001](https://biobank.ctsu.ox.ac.uk/crystal/field.cgi?id=40001); [Field 40002](https://biobank.ctsu.ox.ac.uk/crystal/field.cgi?id=40002)), and self-reported medical condition codes ([Field 20002](https://biobank.ctsu.ox.ac.uk/crystal/field.cgi?id=20002)) reported at the baseline or subsequent UKB assessment centre visit.

For diagnoses that were not listed as “first occurrence” we separately used Primary Care Read V2 codes and ICD-10 codes in Hospital Inpatient data and Death Register records. Information about codes used for exclusion are reported in Supplementary Table 3.

A sensitivity analysis was performed on the leading STR results from the discovery analyses. STRs that reached a *P* < 0.05 were considered statistically significant. A power calculation was also performed to compare the statistical power of the sensitivity analysis with the other two phases of the study. The “pwr.2p.test” function from the “pwr” R package was used.

**SUPPLEMENTARY RESULTS**

**Sensitivity analysis of discovery lead STRs**

We performed sensitivity analysis for the 146 autosomal and chrX 50 + 129 STR alleles (male + female associations) that passed Bonferroni correction in the discovery analysis in 30 brain volumes’ analyses.

Of all significant findings from the discovery phase, 62 STRs autosomal STR regions, 35 STRs (in male analysis) and 69 STRs (in female analysis) in the chromosome X showed significant association in the sensitivity analysis. Of the six STRs in the chromosome X associated with the basal ganglia region (caudate, putamen, pallidum) in either female or male sex groups, only one showed consistent result in the sensitivity analyses in both sex groups. Autosomal and chromosome X sensitivity analyses’ results are reported in Supplementary Table 8-9, respectively.

*A priori* power analysis showed that the smaller cohort used in the sensitivity analysis has less statistical power compared to the main discovery analysis. Results of the power analysis are reported in the Supplementary Table 5.

**Autosomal chromosomes: gene concordance between STR-GWAS and SNP-GWAS results associated to traits other than brain volumes.**

For other non-volumetric brain MRI measurements, we found that 9 STR regions [(a) chr12:105467463-105467489, GT; (b) chr14:57129041-57129077, TTA; (c) chr11:128567611- 128567647, TTTTGT; (d) chr20: 53752297-53752321, GT; (e) chr14:59173671-59173695, AAAT and chr14:59140004-59140022, TG; (f) chr3:115088940- 115088952, AAATAT; (g) chr17:71936313-71936355, GT; (h) chr5:35090582-35090606, TTG; (i) chr8:115856855-115856895, AGAT] were associated to vertex-wise MRI cortical surface area, cortical thickness and/or sulcal depth in previous SNP-based GWAS: (a) the *C12orf75*/*CASC18* locus was associated to the cortical surface area^3,4^; (b) the *OTX2-AS1* and (c) *ETS1* gene regions were associated with cortical surface area^3,4^ and the sulcal depth^4^; (d) the *RNU7-14P* gene region was associated with the cortical surface area^3^ and cortical thickness^4^; (e) the *LINC01500*/*DAAM1* locus was associated with cortical surface area^3-7^, cortical thickness^3-5^ and sulcal depth^4^ ; (f) the *ZBTB20* and (g) the *ROCR* regions were associated with sulcal depth^4^; (h) the *PRLR* locus was associated with cortical surface area^3^; (i) *TRPS1* locus was associated with sulcal depth^4^ and cortical thickness^4^.

We found that the among the STRs identified in our analyses eighteen STR genomic regions are also associated with mental health disorders (such as hyperactivity disorder, autism spectrum disorder, bipolar disorder, major depression, obsessive-compulsive disorder, schizophrenia) (Supplementary Table 11), four with cognitive functions [*NR4A2*/*GDP2* (chr2:156342362- 156342390, CCCCCGG); *DENND5B* (chr12:31443044- 31443064, TG); *ADAMTS5* (chr21:26957447-26957479, GATA*)*; *BIRC6* (chr2:32440751-32440778, TTA)], ten with math ability (Supplementary Table 11), nine with educational attainment (Supplementary Table 11), three with intelligence and verbal-numerical reasoning [*BIRC6* (chr2:32440751-32440778, TTA), *NR4A2*/*GDP2* (chr2:156342362- 156342390, CCCCCGG), *DENND5B* (chr12:31443044- 31443064, TG)], four with neuroticism [*NR4A2* (chr2:156342362- 156342390, CCCCCGG), *OTX2-AS1* (chr14:57129041-57129077, TTA), *SLC44A5* (chr1:75553883-75553913, AGG), *SLC7A8* (chr14:23160944- 23160960, TC), *TRPS1* (chr8:115856855-115856895, AGAT)], two with Parkinson’s disease (*CCDC62/HIP1R* (chr12:122834335-122834365, GGAAA), *ITGA8* (chr10:15697046-15697066, AC)], one with reaction time [*MCC* (chr5:113093469-113093491, TC)], one with multiple sclerosis [*ETS1,* (chr11:128567611- 128567647, TTTTGT)], one with amyotrophic lateral sclerosis [*TIAM1*, (chr21:31280032-31280048, CA)], four with sleep-related traits (*OTX2-AS1* (chr14:57129041-57129077, TTA), *RNU6-472P* (chr12:33779011-33779027, AAAC), *LINC02229* (chr5:66344987-66345007, TG), *KCNMB2* (chr3:178847784-178847808, CT), *NLGN1* (chr3:17388998831740867, TA), *OSBPL1* (chr3:31740857-31740867, TA), *TRPS1* (chr8:115856855-115856895, AGAT)]; and nine with pathological hallmarks in Alzheimer’s disease (Supplementary Table 11). A complete list of results can be found in the Supplementary Table 11.

**X chromosome: gene concordance between STR-GWAS and SNP-GWAS results associated to traits other than brain volumes.**

In the chrX analysis in male participants, the following gene regions were reported in the GWAS-catalog: in the The *PLAC1* gene region (chrX:134560494-134560512, GT) was previously reported in GWAS in association with brain shape^8^; the *GPC3* gene region (chrX:133923131-133923151, AC) was associated with educational attainment^9^; the *IL1RAPL1, EDA* and *GPC3* gene regions (chrX:28996620-28996640, TG; chrX:69674102-69674146, CTAT; chrX:133923131-133923151, AC) were previously associated with sleeping related traits/disorders^10^ (Supplementary Table 12a).

In the chrX analysis in female participants, the following gene regions were reported in the GWAS-catalog: the *DACH2* gene region (chrX:86283871-86283893, AC; chrX:86547524- 86547568, AC) was previously reported in GWAS in association with brain shape^8^; the *BEX1*/*NFX3*, *GPC3* and the DMD gene regions (chrX:103065765-103065795, TCT; chrX:133882535-133882559, TG; chrX:31922495-31922525, GT) were associated with educational attainment^9^; the *GPC3* gene region (chrX:133882535-133882559, TG) was previously associated with sleeping related traits/disorders^10^ (Supplementary Table 12b).

**Supplementary Figure 1: Distribution of catalogues’ STR by repeat unit**

Repeat unit length = repeat unit length group expressed in number of base pairs (bp).

Count = count of each STR repeat unit length group expressed in percentage.

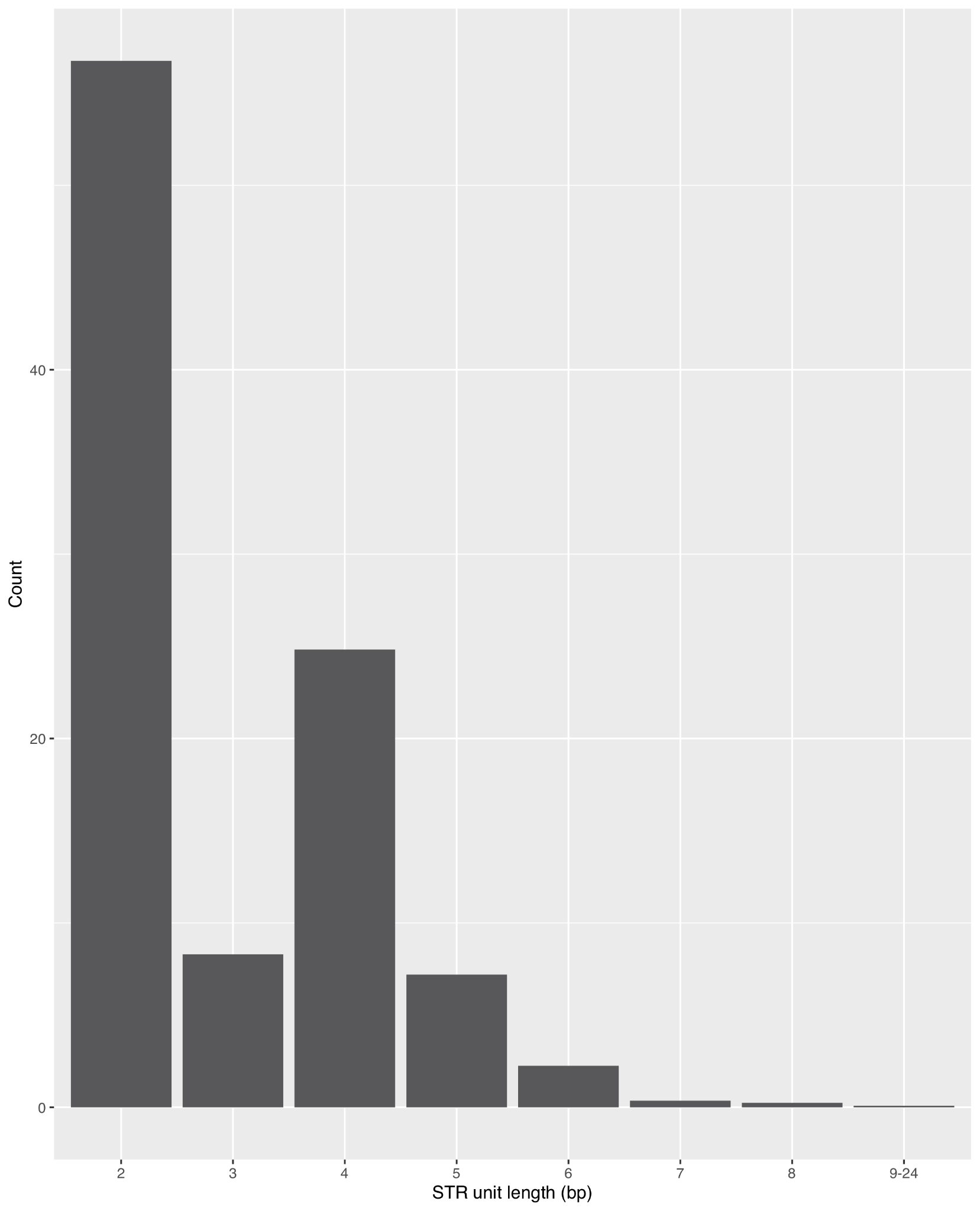
